# Comprehensive Study of Germline Mutations and Double-Hit Events in Esophageal Squamous Cell Cancer

**DOI:** 10.1101/2021.02.04.21251116

**Authors:** Bing Zeng, Peide Huang, Peina Du, Xiaohui Sun, Xuanlin Huang, Xiaodong Fang, Lin Li

**Author notes:** **Correspondence:** Lin Li, Xiaodong Fang.

## Abstract

Esophageal squamous cell cancer (ESCC) is the eighth most common cancer around the world. Several reports have focused on somatic mutations and common germline mutations in ESCC. However, the contributions of pathogenic germline alterations in cancer susceptibility genes (CSGs), highly frequently mutated CSGs, and pathogenically mutated CSG-related pathways in ESCC remain unclear. We obtained data on 571 ESCC cases from public databases and East Asian from the 1000 Genomes Project database and the China Metabolic Analytics Project database to characterize pathogenic mutations. We detected 157 mutations in 75 CSGs, accounting for 25.0% (143/571) of ESCC cases. Six genes had more than five mutations: *TP53* (n = 15 mutations), *GJB2* (n = 8), *BRCA2* (n = 6), *RECQL4* (n = 6), *MUTYH* (n = 6), and *PMS2* (n = 5). Our results identified significant differences in pathogenic germline mutations of *TP53, BRCA2*, and *RECQL4* between the ESCC and control cohorts. Moreover, we identified 84 double-hit events (16 germline/somatic double-hit events and 68 somatic/somatic double-hit events) occurring in 18 tumor suppressor genes from 83 patients. Patients who had ESCC with germline/somatic double-hit events were diagnosed at younger ages than patients with the somatic/somatic double-hit events, although the correlation was not significant. Fanconi anemia was the most enriched pathway of pathogenically mutated CSGs, and it appeared to be a primary pathway for ESCC predisposition. The results of this study identified the underlying roles that pathogenic germline mutations in CSGs play in ESCC pathogenesis; increased our awareness about the genetic basis of ESCC; and provided suggestions for using highly mutated CSGs and double-hit features in the early discovery, prevention, and genetic counseling of ESCC.

## 1 Introduction

Esophageal squamous cell cancer (ESCC) is one of the most common cancers in the world, and it is especially common in Asian countries, North America, and the eastern corridor of Africa (1). In China, there are approximately 478,000 new cases and 375,000 deaths related to ESCC each year (2). Many factors reportedly have relationships with ESCC; these include smoking, drinking, and dietary habits (3). However, the hereditary factors involved in ESCC remain unclear. Thus, understanding the genetic mutations and molecular events in ESCC might be pivotal to reduce the incidence and mortality rate of ESCC.

Enormous efforts have been taken to identify somatic alterations by whole-genome sequencing (WGS) or whole-exome sequencing (WES) (4,5), and several studies reveal the complex process of tumor development (6,7). Many common germline single-nucleotide polymorphisms (SNPs) have been identified by genome-wide association studies (8–16). rs138478634, a *CYP26B1* low-frequency variant, was proved to be involved in ESCC development (14). In 2018, several pan-cancer studies focused on pathogenic germline mutations to explore hereditary factors in cancers; 871 rare cancer predisposition mutations and copy number variations (CNVs) were observed in 8% of 10,389 cases, and 7.6% of the 914 patients with pediatric cancers had tumors that harbored pathogenic mutations in cancer predisposition genes (17,18). Deng et al. (19) identified germline profiles in Chinese patients with ESCC and uncovered the association between genotype and environment interactions.

Additionally, *BRCA2* was associated with ESCC risk in Chinese patients (20). Reflecting a critical part of cancer susceptibility, the two-hit hypothesis assumes that hereditary retinoblastoma involves double mutations and that one mutation is in germline DNA whereas nonhereditary retinoblastoma involves two somatic mutations (21). On the basis of these findings, double-hit events in some studies were used to identify cancer predisposition genes (22,23). These studies demonstrated the significance of pathogenic germline mutations and double-hit events in genetic testing and risk assessment for cancer.

To our knowledge, cancer predisposition genes and molecular events in ESCC remain poorly understood. Here, we identified pathogenic/likely pathogenic germline predisposition mutations and highly frequently mutated CSGs in a large ESCC cohort. We discovered significantly different pathogenic germline mutations of *TP53, BRCA2*, and *RECQL4* in ESCC cohorts, and we clarified the association between double-hit events and diagnosis age in patients with ESCC. In addition, we identified pathogenically mutated CSG-related pathways for ESCC to illuminate the mechanism affected by pathogenic mutations. Results of this study will improve genetic testing for relatives of patients with ESCC and facilitate implementation of organizational or institutional measures for ESCC prevention and surveillance.

## Materials and Methods

### 2.1 Sample acquisition

We collected 592 ESCC samples from published studies and The Cancer Genome Atlas (a total of nine projects) (Supplementary Table 1), and we excluded poor-quality samples and hypermutant samples (4,5,24–29). Clinical information is listed in Supplementary Table 2. The WGS and WES data from the same studies came from distinct patient cases. The quality control analysis uncovered an average sequencing depth of 55×∼161× for WES samples and 30×∼65× for WGS samples (Supplementary Figure 1A), the 10× average coverages were more than 90% in most WES and WGS samples (Supplementary Figure 1B). Moreover, the relationship between 10× average coverages and average sequencing depths showed a positive correlation (Supplementary Figure 1C), suggesting that the qualities of most samples were proofed. The mean depth of our data and the public databases we used as controls were able to provide enough variants to execute downstream analysis (30). The study protocol was reviewed by the institutional review board of the Beijing Genomics Institution.

**Table 1.**
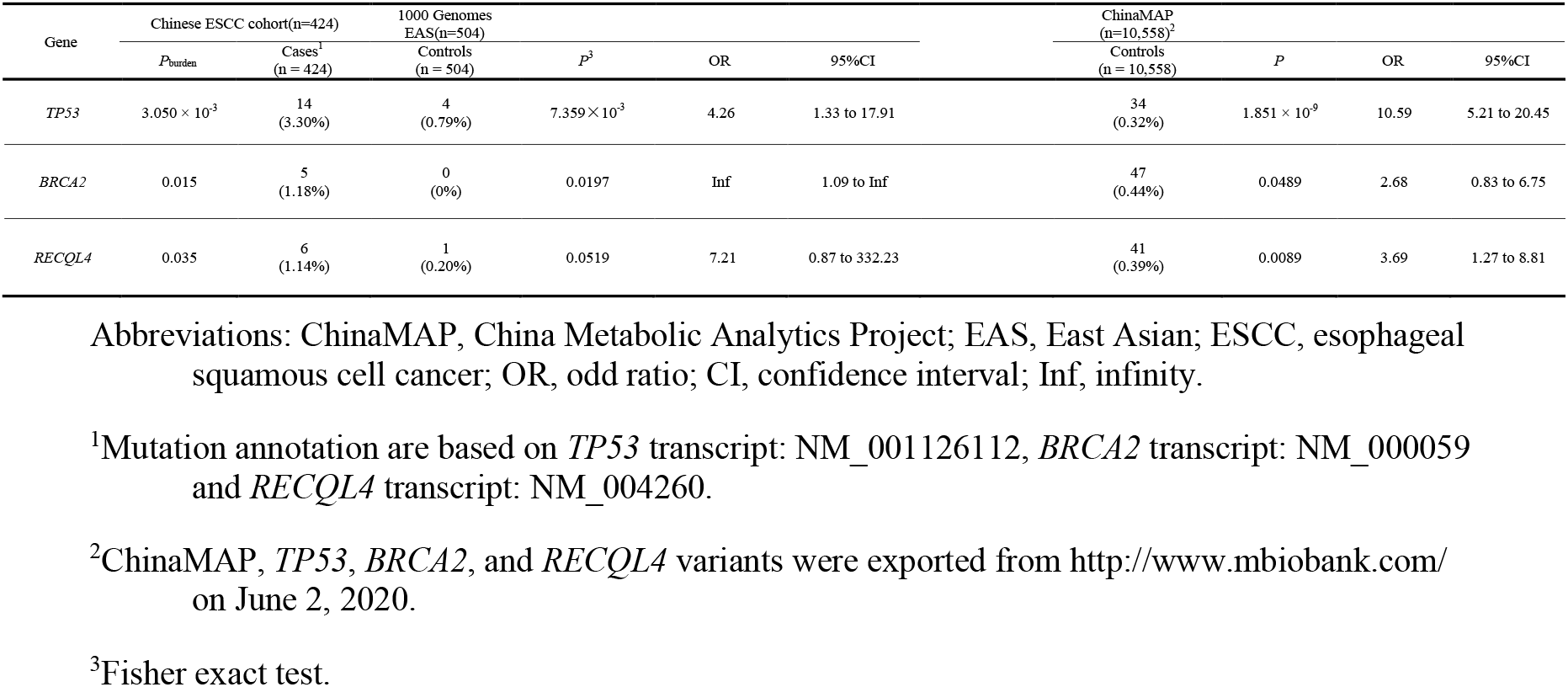
Significance of *TP53, BRCA2*, and *RECQL4* pathogenic or likely pathogenic variants for ESCC risk in Chinese patients

**Figure 1.**
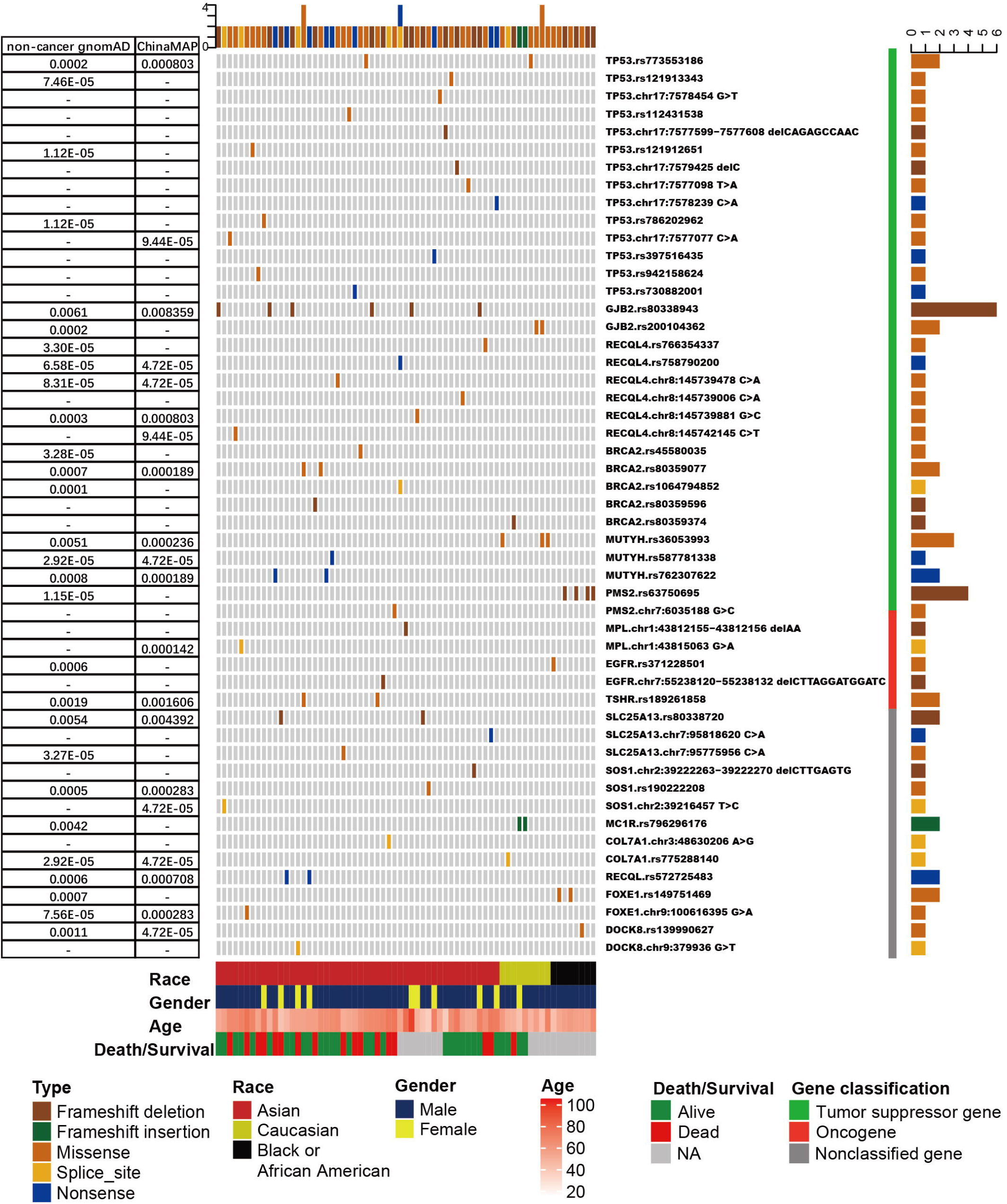
The frequency and distribution of cancer susceptibility genes (CSGs) with more than one pathogenic/likely pathogenic germline mutation detected in patients with esophageal squamous cell cancer (ESCC). Only tumor suppressor genes with more than five mutations are shown. Upper bars represent the cumulative mutation numbers of each sample. Bottom bars represent the clinical information (race, gender, age, and survival/death) about the patients. The left table presents the frequency of mutations shown in the noncancer Genome Aggregation Database (gnomAD) and the China Metabolic Analytics Project (ChinaMAP) database. Right bars represent the mutation counts. The classification of the CSG is next to the mutation name (gene name + reference SNP number or gene name + chromosome position + nucleotide change).

### 2.2 Data processing and mutation calling

The fastq data from 571 samples (38 WGS samples, 533 WES samples) were trimmed and filtered using SOAPnuke (v1.5.6 with default parameters, except where -n 0.1 -l 11 -q 0.5 -G -T 1) (31). Data from ESCC-P006 was transformed from bam files using the GATK SamToFastq (v4.0.6.0 with default parameters) (32). The high-quality reads were aligned to the hg19 human reference genome with a Burrows-Wheeler Aligner (v0.7.17-r1194-dirty with default parameters, except where -o 1 -e 50 -m 100000 -i 15 -q 10 -a 600) (33). MarkDuplicates GATK (version as above with default parameters, except where -CREATE_INDEX true, -reportMemoryStats true, - VALIDATION_STRINGENCY SILENT) was used to mark duplicated reads. BaseRecalibrator (version as above with default parameters) and ApplyBQSR (version as above with default parameters, except where -create-output-bam-index true) were performed to base quality score recalibration (32). Germline variants were joint-called using GenotypeGVCFs (version as above with default parameters, except where -ignore-variants-starting-outside-interval true) after CombineGVCFs (version as above with default parameters) and annotated with the Variant Effect Predictor (VEP v98.3) (32,34). The calling germline variants of nine projects are shown in Supplementary Figure 1D. Samples with fewer than 80,000 variants were filtered out. Somatic variants were detected by GATK MuTect2 (version as above with default parameters except where - af-of-alleles-not-in-resource 0.0000025, -native-pair-hmm-threads 1, -add-output-vcf-command-line false), and Oncotator (v1.9.9.0) was used for annotation (32,35). Loss of heterozygosity (LOH) and other somatic CNVs (SCNVs) were detected with FACETS (v0.5.14) and Pathwork (v1.0) for 533 WES and 38 WGS samples, respectively (36,37).

### 2.3 CSG sets

We curated CSGs from published papers and the Catalogue of Somatic Mutations in Cancer (COSMIC) database (38); we included cancer predisposition genes from three papers (17,18,39) and genes with recorded germline associations in COSMIC (Supplementary Table 4). After we removed duplicated genes, the CSG set included 260 genes. CSGs were divided into three groups according to the literature (17,40–42); these groups were tumor suppressor genes (TSGs; n = 139), oncogenes (n = 36), and nonclassified genes (n = 85).

### 2.4 Pathogenicity evaluation

We first leveraged an in-house pathogenicity database to match germline variants; the rest of the germline variants were evaluated using InterVar (InterVar_20190327) as a supplemental method to find germline pathogenic/likely pathogenic mutations (43). Germline pathogenic or likely pathogenic variants are hereafter referred to as pathogenic mutations. The pathogenicity database included ClinVar, the Human Gene Mutation Database, mutations collected from papers, and mutations we assessed according to consensus guidelines by the American College of Medical Genetics and Genomics and the Association for Molecular Pathology (17,44–46). We filtered for pathogenic variants with an allele frequency of 0.5% or lower in the Genome Aggregation Database (gnomAD version v2.1) (47). Pathogenic mutations in 260 high-interest CSGs (Supplementary Table 6) were selected for analysis and were checked by Deep Variant (48); manual verification ruled out false-positive results. For somatic nonsilent variants, with the exception of frameshift, nonsense, and splice-site mutations, three silico tools—SIFT (49), Polyphen2_HDIV (50) and CADD (51) were used to predict pathogenicity. If a variant was predicted as damaging in any two silico tools (SIFT: D, Polyphen2_HDIV: D/P, CADD score > 15), the variant was categorized as deleterious (39,52).

### 2.5 Identification of potential double-hit events

According to the two-hit hypothesis, potential double-hit events are identified after two or more hits have been found in the same CSG; in this study, we set rigorous standards for determining hits.

Pathogenic germline mutations were considered hits. Effective somatic variations were defined as hits if they met the following requirements: frameshift, nonsense, splice-site mutations, or deleterious missense and in-frame variants and SCNVs that caused allele loss. Copy-neutral LOH, duplication LOH, homozygous deletion, and hemizygous deletion were assumed to be linked to allele loss and were termed allele loss SCNVs (53,54). Integrative Genomics Viewer software was used to examine the authenticity of biallelic events (55). For double-hit events comprised of germline hits and allele loss SCNVs, we calculated SNP average depths and variant allelic frequency in normal and tumor tissues of ESCC to further validate allele loss SCNV events. Samples with variant allele frequencies less than 0.5 in tumors were removed.

### 2.6 Statistical analyses

To evaluate the correlations of the clinical features and genetic events, we used the two-sided Student’s t-test. We conducted the two-sided Fisher’s exact test to assess the gene-based association analysis and pathway enrichment. We also performed a burden test to determine the exact relationships between pathogenic mutations in CSGs and ESCC (56); *p* < 0.05 was defined as statistically significant.

## 3 Results

### 3.1 Population characteristics

Overall, 469 of 571 patient cases were Asian (424 Chinese, 41 Vietnamese, one Canadian, one Brazilian, two without country information), 41 were Caucasian, 58 were Black or African American, and the rest were Brazilian without ethnicity information. The entire population consisted of 105 women, 465 men, and one patient without gender information. The average diagnosed age for 567 patients (the rest had no information) was 58.81 years (the minimum diagnosed age was 24 years, and the maximum diagnosed age was 93 years). Thirty-five patients had family histories of ESCC, and the average age of patients with ESCC with a family history (mean age and standard deviation [SD] was 56.80 [9.3] years; [range, 41–82 years]). This average was lower than the age of patients with ESCC without a family history (mean age [SD], 60.00 [8.2] years; [range, 36–78 years]; t-test *p* = 0.059; 95% CI, -6.511 to 0.121) (Supplementary Figure 2). The average survival for 399 patients (the rest had no information) was 879.8 days (minimum survival, 3 days; maximum survival, 2,580 days). In this study, 347 patients had a smoking history, and 215 patients had histories of alcoholism. With regard to disease grade, 334 patients had disease with pathological grade 2 or lower, and 86 patients had disease with pathological grade greater than 2; pathological grade information was missing for 151 patients. All patients were diagnosed with disease stages I (n = 72), stage II (n = 207), stage III (n = 203), and stage IV (n = 7); 82 patients were not assigned disease stages for this study (their information was lost).

**Figure 2.**
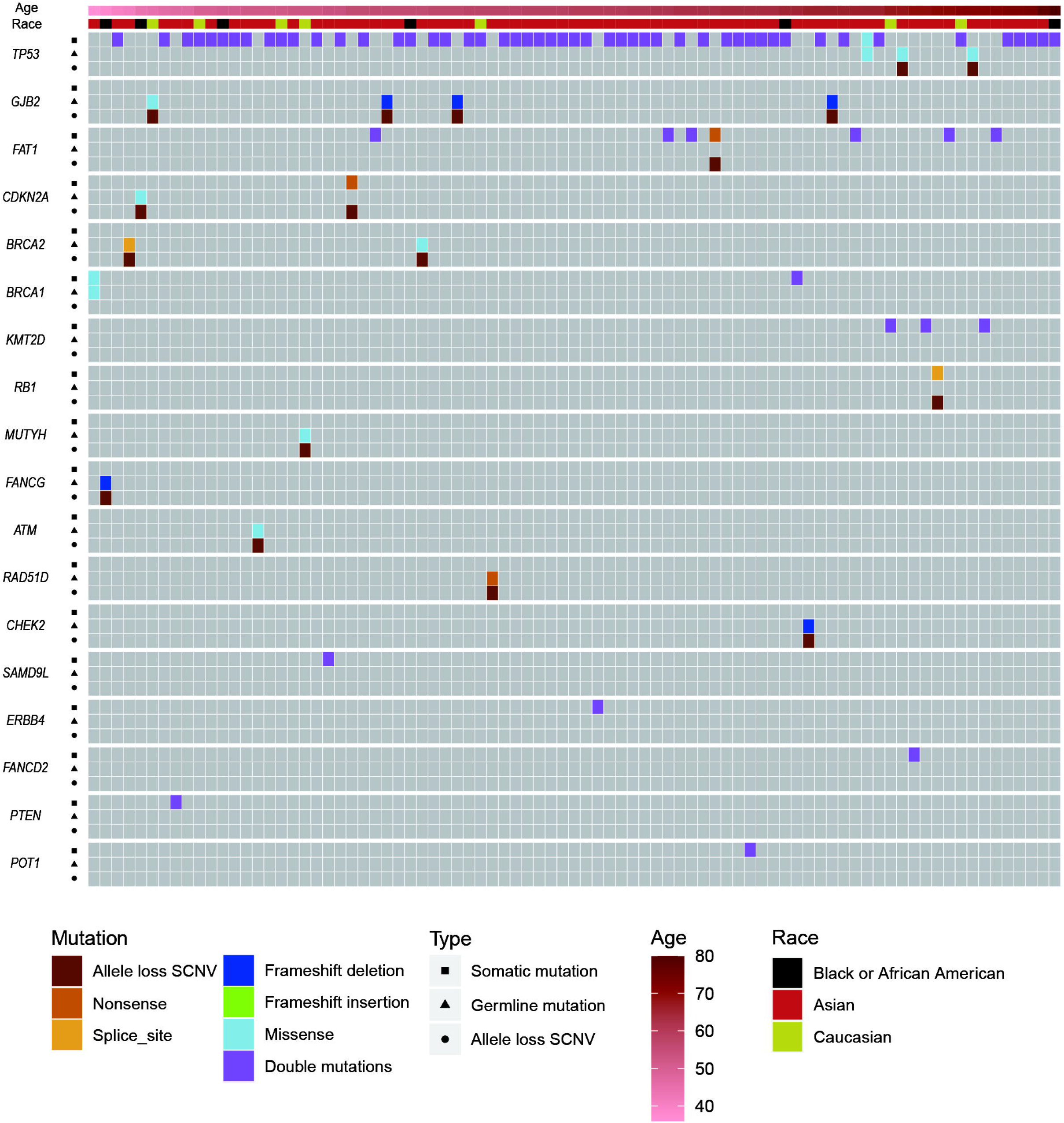
The distribution of pathogenic/likely pathogenic germline mutations, somatic mutations, and allele loss somatic copy number variations (SCNVs) in esophageal squamous cell cancer (ESCC) cases with potential double-hit events. Upper bars represent the clinical information (age and race) about those patients. Squares represent somatic mutations, triangles represent germline mutations, and circles represent allele loss SCNVs.

### 3.2 Pathogenic germline mutations in CSGs

Overall, 2,484 pathogenic germline mutations were identified, including 1,973 SNPs and 511 insertions or deletions (Supplementary Table 5). Each sample had an average of 4.4 pathogenic mutations. After filtration by CSGs, 157 pathogenic mutations (113 SNPs and 44 insertions or deletions) were discovered from 25.0% (143/571) of the population (Supplementary Figure 3). Although each sample had an average of 1.1 pathogenic mutation in CSGs, only 12 (2.10%) of the 571 patients harbored one or more pathogenic mutation in CSGs (Figure 1, Supplementary Table 6). The frequency of most mutations was rare in the gnomAD noncancer database and in the China Metabolic Analytics Project (ChinaMAP) database (47,57), indicating the sparsity of these deleterious mutations in the general population. As expected, most of the frequently mutated CSGs belonged to TSGs, and they were involved in biological processes, such as DNA repair.

**Figure 3.**
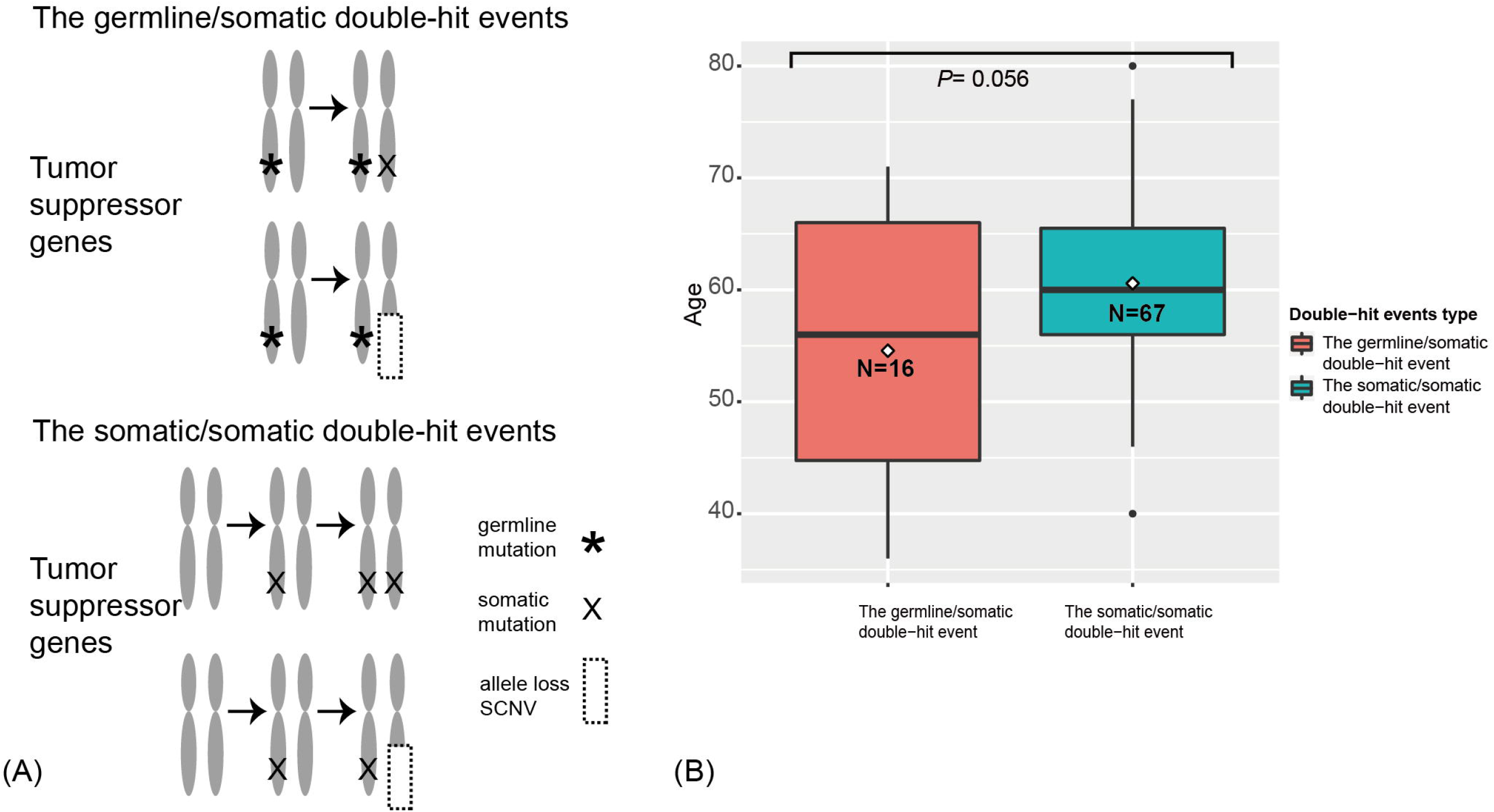
The two types of double-hit events. **(A)** The paradigm of double-hit events. **(B)** The correlation between age and double-hit event type in esophageal squamous cell cancer (ESCC) cases. The position of line is the median age, and the position of rhombus is the mean age in specific ESCC cohorts. The digits in the boxes are the numbers of ESCC cases in each category.

In general, the CSGs detected more than five times were *TP53* (n = 15 mutations), *GJB2* (n = 8), *BRCA2* (n = 6), *RECQL4* (n = 6), *MUTYH* (n = 6), and *PMS2* (n = 5). *TP53* was the most frequently mutated CSG, with pathogenic germline mutations in 2.63% (15/571) of patients with ESCC (Figure 1, Supplementary Table 6 and Supplementary Figure 4); The result was the same as *TP53* pathogenic mutations in a study of osteosarcoma (39). In our study, 86.7% (13/15) of *TP53* mutations were nonsynonymous single-nucleotide variations. c.A1073T (rs773553186; in 0.35%, or 2/571) and c.C742T (rs121912851; in 0.18%, or 1/571) were recorded in the International Agency for Research on Cancer TP53 database (58). All *TP53* pathogenic mutations were found in Chinese patients, except c.A1073T (one each in a Chinese and a Caucasian patient) (Supplementary Figure 4). Three of the *TP53* mutations, c.C742T, c.C586T, and c.C817T, have been reported in osteosarcoma (39), and *TP53* c.C742T has also been identified in low-grade glioma (17) (Supplementary Figure 4).

**Figure 4.**
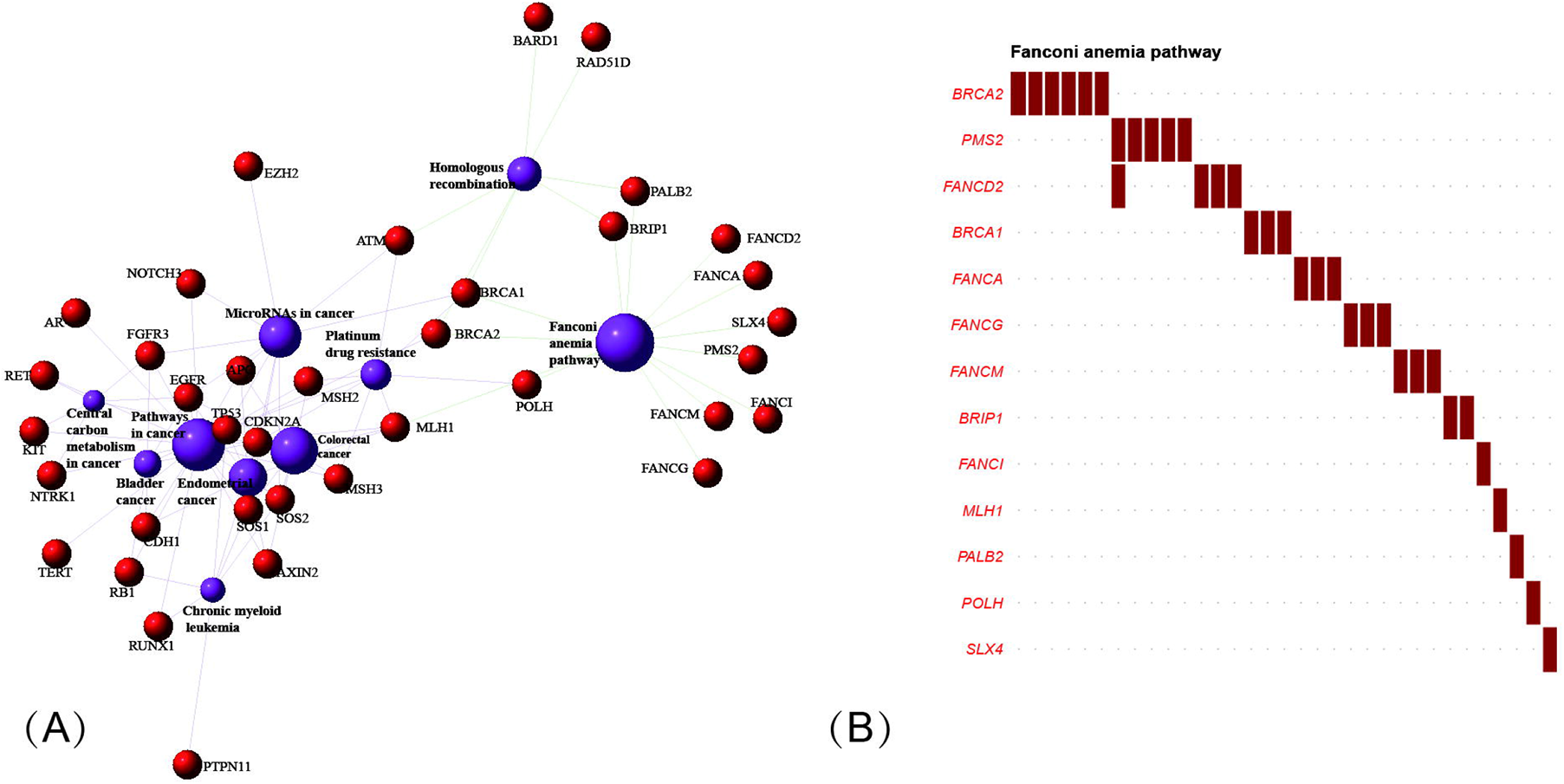
Significantly enriched pathways and networks in esophageal squamous cell cancer (ESCC). **(A)** The network composed of genes involved in the top 10 pathways in the Kyoto Encyclopedia of Genes and Genomes pathway enrichment. The red dots represent genes, and the blue circles represent pathways. The larger the area, the higher the degree of enrichment. The different lines represent various categories of pathways; green lines indicate genetic information processing, and purple lines indicate human disease. Solid-line rectangles and solid rectangles in blue, red, and yellow represent various gene classifications. **(B)** The x axis represents cancer susceptibility genes mutated in the Fanconi anemia pathway; the y axis represents the number of patients affected in our cohort. Red font: tumor-suppressor genes.

Pathogenic mutations in *GJB2* represented the second most frequently mutated CSGs (Figure 1); their detection rate was 1.40% (8/571). The c.235delC (rs80338943) mutation, a common pathogenic frameshift deletion mutation in East Asian (EAS) populations, has been detected in six Asian (Chinese) patients with ESCC (59). Because this mutation has not been detected in other populations, rs80338943 may be specific to Chinese or Asian populations.

Nonsynonymous single-nucleotide variations occupied no less than 50% of pathogenic germline mutations in *BRCA2, RECQL4*, and *MUTYH* (Supplementary Table 6). In the upstream region, we detected a pathogenic splice mutation, *BRCA2* c.-39-1_-39delGA (rs758732038), in a patient, and the mutation was reported in ClinVar as likely pathogenic (46). The mutation has also been reported in patients with breast cancer and medulloblastoma (60–62). *RECQL4* pathogenic mutations were only detected in Asian (Chinese) patients in our study, and *RECQL4* c.C2272T has been reported in ovarian cancer/Rothmund–Thomson syndrome. In our study, *MUTYH* c.C1178T (rs36053993) and c.C458T (rs762307622) were detected three times (0.53%, or 3/571) and two times (0.35%, or 2/571), respectively. rs36053993 only detected in Caucasian patients and rs762307622 only detected in Asian (Chinese) patients. From gnomAD, rs36053993 in a homozygous state was found in three non-Finnish Europeans; this mutation may have been caused by founder events (63,64). Pathogenic mutations in *PMS2* were detected five times in five patients in our study (0.88%), and c.2192_2196delAGTTA (rs63750695) was observed in only four patients, who were all African. The rs63750695 mutation has also been discovered in Lynch syndrome, colorectal cancer, and ovarian carcinoma (65–67); however, it was rare in noncancer gnomAD and ChinaMAP, for which frequencies were 1.15 × 10^−5^ and 0, respectively (Figure 1). rs63750695 is possibly specific to African ethnicity in ESCC.

The total number of pathogenic germline mutations and the frequency of mutations were relatively lower in oncogenes and nonclassified genes compared with TSGs. *TSHR* and *MPL* were oncogenes that were mutated in two patients with ESCC; other oncogenes occurred in just one patient. *SLC25A13* was one of the nonclassified genes with the most pathogenic mutations.

We also investigated our pathogenic germline mutations in a previous pan-cancer study (17). Nine mutations were spread over 22 samples with diverse cancers (Supplementary Table 9). *SLC25A13* c.852_855delCATA (n = 7), *GJB2* c.235delC (n = 7), and *PALB2* c.C2257T (n = 2) were the variants observed more than once across cancers. We detected multiple susceptibility loci (31/47), also identified in previous genome-wide association studies, in our patients with ESCC (Supplementary Table 10) (8–16). Of those genes with susceptibility loci, pathogenic mutations *PDE4D* c.T108A and *RUNX1* c.61+1delG were found in two patients separately (Supplementary Table 5). We also confirmed from the COSMIC database that 87.3% (137/157) of pathogenic mutations in CSGs had nonsilent somatic mutations in the same or a nearby (within five) amino acid position (Supplementary Table 6). Among 137 mutations, 107 mutations were observed in TSGs, representing 89.2% (107/120) of all mutations.

### 3.3 Pathogenic germline mutations frequency in ESCC cases versus controls

To reveal the relationships between highly frequent mutated CSGs and ESCC, we chose the Chinese patients to continue the study, to leverage the most population data and avoid any ethnicity-specific effect. We conducted gene-based association analyses by comparing various germline mutation data from individuals with ESCC versus a 1000 Genomes Project EAS population and versus a ChinaMAP population separately (57,68). We also conducted rare variant burden tests on the ESCC individuals and the 1000 Genomes Project EAS population (68). Through the same pathogenicity evaluation pipeline, pathogenic mutations were identified in two public database populations.

Analysis of results identified significantly higher pathogenic mutations in Chinese patients with ESCC versus public population databases (including 1000 Genomes Project EAS and ChinaMAP data), as reflected by odd ratios (ORs) of pathogenic mutations in *TP53* from the Chinese ESCC populations compared with the 1000 Genomes Project EAS populations (OR = 4.26; 95% CI, 1.33-17.91; Fisher’s exact test *p* = 7.359 × 10^−3^) and compared with the ChinaMAP populations(OR = 10.59; 95% CI, 5.21-20.45; Fisher’s exact test *p* = 1.851 × 10^−9^); in *BRCA2* from the Chinese ESCC populations compared with the 1000 Genomes Project EAS populations (OR= infinity; 95% CI, 1.09-infinity; Fisher’s exact test *p* = 0.0197) and compared with the ChinaMAP populations (OR = 2.68; 95% CI, 0.83-6.75; Fisher’s exact test *p* = 0.0489); and in *RECQL4* from the Chinese ESCC populations compared with the 1000 Genomes Project EAS populations (OR = 7.21; 95% CI, 0.87-332.23l; Fisher’s exact test *p* = 0.0519) and compared with the ChinaMAP populations (OR = 3.69; 95% CI, 1.27-8.81; Fisher’s exact test *p* = 0.0089)(Table 1). Likewise, in the burden analyses (Table 1), the numbers of pathogenic mutations from *TP53* (14/424, or 3.30%; burden test *p* = 3.050 × 10^−3^), *BRCA2* (5/424, or 1.18%; burden test *p* = 0.015), and *RECQL4* (6/424, or 1.14%; burden test *p* = 0.035) in our Chinese ESCC cohort were higher than those observed in the 1000 Genomes Project EAS group.

### 3.4 Potential double-hit events

To further survey the genetic predisposition of ESCC, we tried to identify potential double-hit events in ESCC. First, we identified 49,876 nonsilent mutations (Supplementary Table 3) in protein-coding regions from patients with ESCC. (We filtered the somatic mutations that overlapped with our own panel of normal datasets and the Exome Aggregation Consortium database.) Then, by integrating pathogenic germline mutations and effective somatic mutations (Supplementary Table 8) or allele loss SCNVs, we found 84 potential double-hit events (Figure 2). To distinguish hits with germline mutations, the double-hit events were classified as germline/somatic double-hit events and somatic/somatic double-hit events. We identified 16 potential germline/somatic double-hit events (two germline mutations coupled with somatic mutations, and 14 germline mutations accompanied with allele loss SCNVs) (Figure 2, Supplementary Table 11 and Supplementary Figures 5, 6) in 16 patients with ESCC, and we identified 68 potential somatic/somatic double-hit events (three somatic mutations accompanied by allele loss SCNVs and 65 double somatic mutations) (Figure 2, Supplementary Table 12) in 67 cases. The likelihood of two or more somatic mutations happening on the same chromosome was very low (52,69,70). Therefore, we assumed that double somatic mutations were likely in the trans position. Briefly, 83 individuals with ESCC possessed potential double-hit events, representing 14.5% of the ESCC cohort (Figure 2). Notably, one patient had two somatic/somatic double-hit events in different genes.

*GJB2* and *TP53* were the top two CSGs that found germline/somatic double-hit events. Germline/somatic double-hit events were identified in eight CSGs, including *BRCA2, BRCA1, MUTYH, CDKN2A*, and *ATM*. The dominant type of germline/somatic double-hit events was a germline mutation accompanied by an allele loss SCNV. In the remaining, germline mutations were coupled with somatic mutations; these were only discovered in *TP53* and *BRCA1*, possibly because SCNVs are relatively abundant in tumors and cover large genome region. In the somatic/somatic double-hit events, the *TP53* gene had the highest frequency, and most of the remaining genes had one potential double-hit event. Double somatic mutation was the main type of somatic/somatic double-hit event (Supplementary Table 12).

When we compared diagnosis ages of patients with different double-hit events, we found that patients with germline/somatic double-hit events (with pathogenic germline mutations) had younger diagnosis ages (mean age [SD], 54.6 [11.2] years; [range, 36–71 years]) compared with patients in the somatic/somatic double-hit events (without pathogenic germline mutations; mean age [SD], 60.6 [7.8] years; [range, 4–80 years]; t-test *p* = 0.056; 95% CI, -12.216 to 0.177) (Figure 3). The comparison was nonsignificant, maybe it’s due to the limited number of samples with double-hit events in this comparison. However, the finding was consistent in the study by Knudson (21). Using the empirical cumulative distribution function (ecdf) to calculated the expression percentiles of TSGs in an ESCC-P006 cancer cohort, two patients with somatic/somatic double-hit events showed low expression: one in *TP53* (5.32%) and one in *PTEN* (6.38%) (Supplementary Figure 8) (17). Those results support the two-hit hypothesis and suggest that genetic screening in specific TSGs can detect patients with germline/somatic double-hit events earlier.

### 3.5 Pathway enrichment

To obtain a more comprehensive understanding of pathogenic germline genetic mutations affecting pathways, Kyoto Encyclopedia of Genes and Genomes pathway enrichment analyses were performed for multiple gene lists. The Fanconi anemia (FA) pathway was the most significantly enriched in the analysis of 75 pathogenically mutated CSGs (Fisher’s exact test *p* = 6.634×10 ^-19^) (Figure 4A, Supplementary Table 7). In addition, 1,226 pathogenic mutated genes and the genes involved in germline/somatic double-hit events were significantly enriched in this pathway. The top four pathways for CSGs involved in somatic/somatic double-hit events versus for CSGs involved in germline/somatic double-hit events differed significantly (Supplementary Figures 7A-D).

In the tumor-suppressor network, the FA pathway functions to preserve genomic integrity by repairing DNA interstrand crosslinks, regulating cytokinesis, and mitigating replication stress (71,72). 33 ESCC samples carried pathogenic mutations in 13 CSGs included in the FA pathway (Figure 4B). The homologous recombination pathway and the mismatch repair pathway described in a previous ESCC project, and associated with cancer susceptibility, were found in our study (Supplementary Figure 7A) (19,73–75). Those pathways were also reported in pathway enrichments of ovarian cancer and osteosarcoma (39,76). We also interrogated the oncogenic signaling pathways upon which our mutated CSGs converged (77). The cell cycle pathway was the most enriched, followed by p53 pathway, the phosphatidylinositol 3’-kinase-Akt pathway, and the receptor tyrosine kinases-Ras pathway.

## 4 Discussion

We reported the profile of pathogenic germline mutations of a larger ESCC cohort comparing with previous studies (17,19). We found 157 pathogenic mutations in CSGs from 143 (25.0%) of 571 patients with ESCC and identified 84 double-hit events in 83 individuals (14.5%). Double-hit events were found in almost all projects in our study except ESCC-P008, which demonstrated that double-hit events are relatively common in ESCC. As far as we know, there was no report about pathogenic mutations in *GJB2, RECQL4, MUTYH* and *PMS2* in ESCC, however, they were discovered in our study. Overall, *TP53, GJB2, BRCA2, RECQL4, MUTYH*, and *PMS2* were highly frequently mutated CSGs. Significant pathways were identified for different CSGs with pathogenic mutations; the FA pathway appeared to be a primary pathway for cancer predisposition in ESCC. We showed that significantly more pathogenic mutations from *TP53, BRCA2* and *RECQL4* occurred in patients with ESCC than in control cohorts, which indicates that these three CSGs may play vital roles in ESCC. Interestingly, *TP53* and *RECQL4* have also been found significantly associated with osteosarcoma (39). The relationship with diagnosis age was not significant in our study, but double-hit events may be pivotal in ESCC carcinogenesis.

We found that *TP53* had the highest frequency of pathogenic germline mutations and the most double-hit events in CSGs. In our study, 80% (12/15) of germline mutations in *TP53* were located in the p53 domain, which functions in DNA binding. This domain contains four conserved regions that are enriched for somatic mutation hot spots and are essential for the function of the TP53 protein as a transcription factor (78,79). Six of the 12 mutations were discovered in conserved regions.

Environmental factors and specific DNA sequences drive higher mutation rates, which may explain why p53 domain was a hot-spot region (80). Those pathogenic *TP53* mutations may disrupt the p53 transcriptional pathway, which would enhance tumor progression and metastatic potential (81). The US Food and Drug Administration had approved drugs against the pocket in p53 domain (82). These drugs provide treatment options to patients with tumors that have mutations in the p53 domain.

Results of studies in other cancers contrast with our findings about *TP53*. In a renal cell carcinoma study, *FH*, instead of *TP53*, harbored the most double-hit events, and *BRCA1* harbored the most in a pan-cancer study(17,22). Previous studies have reported that most double-hit events with *TP53* involve a mutation accompanied by LOH (83,84). However, in our research, double somatic mutations were the dominant type of double-hit event. It was partially due to the lack of researches on *TP53* double somatic mutations before.

*BRCA2* and *RECQL4* harbored more pathogenic germline mutations in ESCC than in public population. *BRCA2* is known for its involvement in breast cancer and ovarian cancer via the homologous recombination pathway, which is essential for repairing damaged DNA (85,86). And studies have reported *BRCA2* mutations related to ESCC risk in Chinese and Turkmen populations (20,87,88). The double-hit events detected in *BRCA2* in our study were germline/somatic double-hit events; the germline mutations were accompanied by allele loss SCNVs. These results were distinct from those reported in pancreatic acinar-cell carcinomas (89). *RECQL4* is a TSG that encodes RECQL4 helicase, which is involved in DNA replication and DNA repair. Germline mutations in *RECQL4* can cause Rothmund-Thomson syndrome and sporadic breast cancer (90). Although the pathogenic mutations in our ESCC cohort and in the 1000 Genomes EAS group were not significantly different (Fisher’s exact test *p* = 0.0519), the difference between them was also confirmed by analysis of the ChinaMAP cohort (Fisher’s exact test *p* = 0.0089). Importantly, this is the first report, to our knowledge, that illustrates the role of pathogenic mutations in *RECQL4* in ESCC.

The PMS2 protein is a homolog of the PMS1 protein (91) and both of them are components of the mismatch repair system. Common polymorphisms of *PMS1* have been positively associated with ESCC in an African population (92). This finding, together with the connection between PMS1 and PMS2, suggests a possible relationship between *PMS2* and ESCC. Double-hit events of mismatch repair genes could result in Lynch syndrome, as described in several studies (70,93), but we did not detect double-hit events in *PMS2* in our ESCC cohort. A larger ESCC cohort study might uncover double-hit events in *PMS2*, which would strengthen our understanding about ESCC susceptibility.

The genetic variations in ESCC are complicated. Although not all ESCC samples carried pathogenic germline mutations in CSGs, the detection rate of pathogenic mutations was close to that found in osteosarcoma (39). Because numerous susceptibility loci reported in genome-wide association studies were found in this research, we acknowledge that pathogenic mutations and known susceptibility loci may inform a genetic basis of ESCC. Our findings of variants and genes shared between ESCC and other cancers suggests that common hereditary factors exist in pan-cancer. Given the interplay of common SNPs and pathogenic mutations reported in breast cancer and colorectal cancer, the interaction between susceptibility loci and pathogenic mutations in ESCC suggests a need for future exploration (94).

To better understand the genetic factors causing ESCC initiation and development, we confirmed the putative germline-somatic interplay by COSMIC proximity match. The results not only support the pathogenicity of those germline mutations but also imply a signal functional relevance between germline and somatic mutations (76). In addition, we identified potential double-hit events in 83 patients with ESCC; though the difference was not significant, the patients with germline/somatic double-hit events were more likely to be diagnosed at younger ages. It is possible that pathogenic mutations confer the earliest genetic hits to TSGs in cells, so a somatic hit alone would cause loss of function in TSGs (95). As a result of double-hit events, the cells generate malignancy. Furthermore, enriched pathways revealed the process of pathogenic mutations that affect ESCC tumorigenesis and development. In patients without pathogenic mutations or double-hit events, limited CSG sets, potential alternations in methylations of a promoter region, germline CNVs, and gene-environmental or gene-lifestyle interactions are possible explanations for ESCC development.

Despite our findings about the genetic characterization of and double-hit events in ESCC, we still acknowledge limitations to our study. The first is our inability to obtain detailed clinical information because of limited access to public databases. Second, merging different data, such as WGS and WES, may induce biases in cohort-wide variant processing. Third, directly adopting variants from different sources may influence comparisons, because the different sources applied distinct platforms and variant detection pipelines. Fourth, our sample size was not large enough for statistical tests, especially for individual variants.

In sum, we report that approximately 25.0% of patients with ESCC harbored at least one pathogenic germline mutation in CSGs, and approximately 14.5% of ESCC cases could be explained by a two-hit hypothesis. Significantly enriched pathways also validated the significance of those pathogenic mutations. Myriad genome variations occur in patients; our findings represent, to our knowledge, the largest discovery of rare, germline predisposition mutations in ESCC so far. These results strengthen the understanding about genetic factors involved in ESCC and will help improve prevention, early detection, and risk management of ESCC for patients. We acknowledge the shortcomings in the analytical methods and the data sources used. Additional studies are needed to improve our observations and results.

## Supporting information

Quality control of normal and tumor tissues in ESCC patients and germline variants

Correlation of age and family history in ESCC samples

The progress of how to select germline pathogenic variants

The lollipop plot of pathogenic or likely pathogenic germline mutatinos in TP53 from 571 ESCC patients

The IGV screens showing germline mutations coupled with somatic mutations in ESCC samples

The scatter plots showing germline mutations accompanied with allele loss SCNVs in normal tissues and tumor tissues of ESCC samples

The top 20 pathways of enrichment of four gene lists

The expression percentiles of TP53 and PTEN in ESCC-P006 cancer cohort

## Data Availability

The raw data of this project can be found in the Sequence Read Archive hosted by the National Center for Biotechnology Information under accession numbers PRJNA315775, PRJNA399748, PRJNA401209, PRJNA230271, PRJNA317404, SRA112617 and SRP034680 and in the European Genome-Phenome Archive under accession number EGAD00001000845.The whole-exome sequencing data of esophageal squamous cell cancer samples from The Cancer Genome Atlas are available from the National Cancer Institute Genomic Data Commons (https://portal.gdc.cancer.gov/). All relevant datasets for this study are available from the authors.

## 5 Data availability statement

The raw data of this project can be found in the Sequence Read Archive hosted by the National Center for Biotechnology Information under accession numbers PRJNA315775, PRJNA399748, PRJNA401209, PRJNA230271, PRJNA317404, SRA112617 and SRP034680 and in the European

Genome-Phenome Archive under accession number EGAD00001000845.The whole-exome sequencing data of esophageal squamous cell cancer samples from The Cancer Genome Atlas are available from the National Cancer Institute Genomic Data Commons (https://portal.gdc.cancer.gov/). All relevant datasets for this study are available from the authors.

## 7 Author Contributions

LL and BZ: conceptualization. BZ: writing manuscript and performing analysis. PD, XS and XH: providing help in analysis. BZ, LL, XH and PD: collected the data from published literature or database. P H revised the manuscript. LL and XF supervised and supported this project.

## 9 Acknowledgments

This study makes use of data generated by the Molecular Oncology Laboratory of Prof. Qimin Zhan, the Translational Medicine Research Center, Shanxi Medical University of Prof. Yongping Cui, the Department of Radiation Oncology, Fudan University Shanghai Cancer Center of Prof. Kuaile Zhao, the Department of Bioinformatics and Computational Biology, The University of Texas MD Anderson Cancer Center of Prof. Han Liang, The Lineberger Comprehensive Cancer Center, University of North Carolina School of Medicine, Prof. Norman E. Sharpless, the Institute of Clinical Pathology, Shantou University Medical College, Prof. Min Su, the Cedars-Sinai Medical Center, UCLA School of Medicine, Prof. H. Phillip Koeffler and Prof. Jie He of Cancer Institute and Hospital Chinese Academy of Medical Sciences. We also acknowledge other Professors for sharing the fastq data, The National Center for Biotechnology Information, The European Genome-phenome Archive, The Cancer Genome Atlas for sharing the esophageal squamous cell cancer data.

The last but not the least, I want to thanks to my wife Panhong Liu, without her support, without my scientific research.

