## Supplementary figures and images for "Comprehensive Study of Germline Mutations and Double-Hit Events in Esophageal Squamous Cell Cancer"

### Correlation of age and family history in ESCC samples

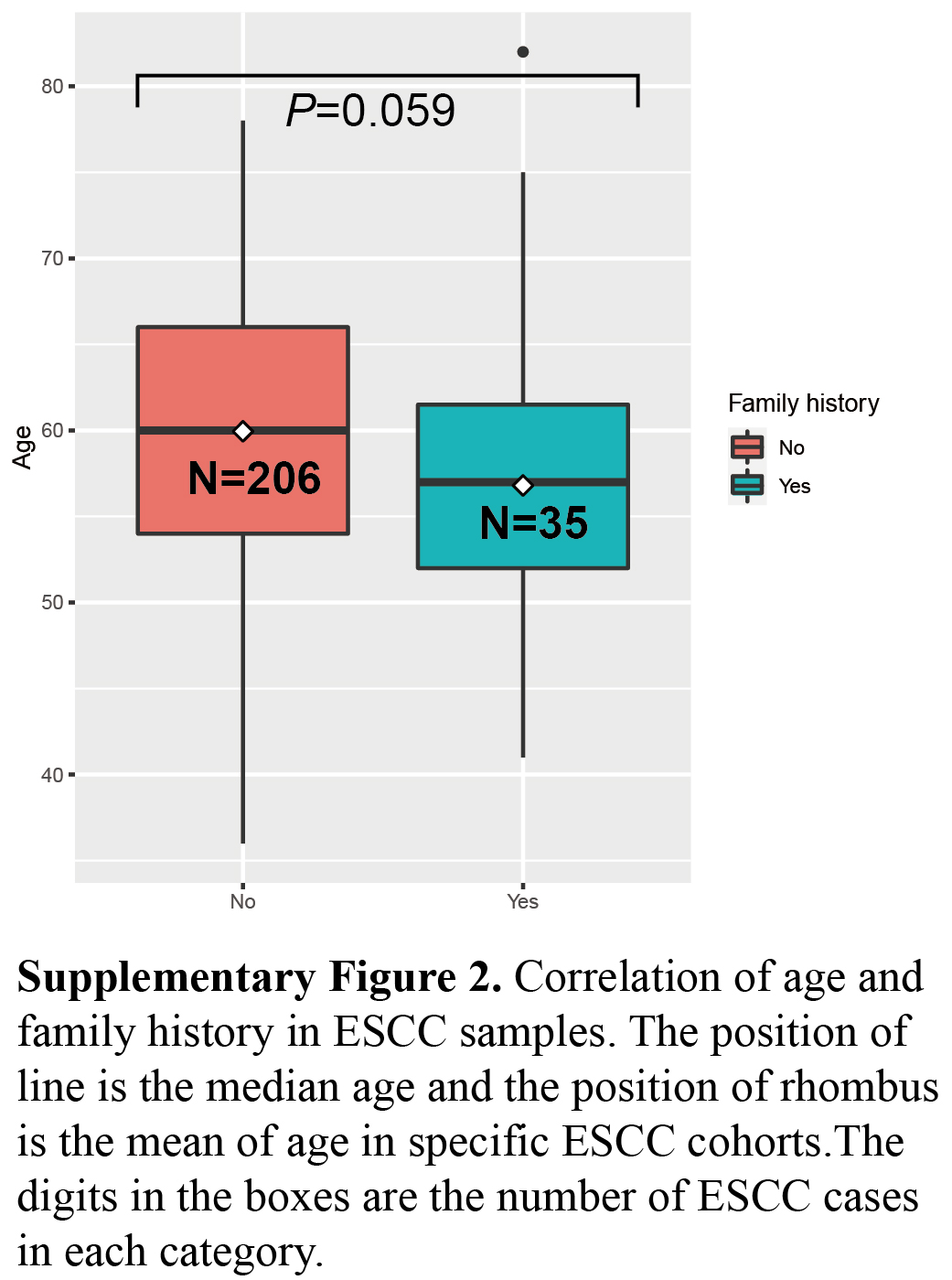

### Quality control of normal and tumor tissues in ESCC patients and germline variants

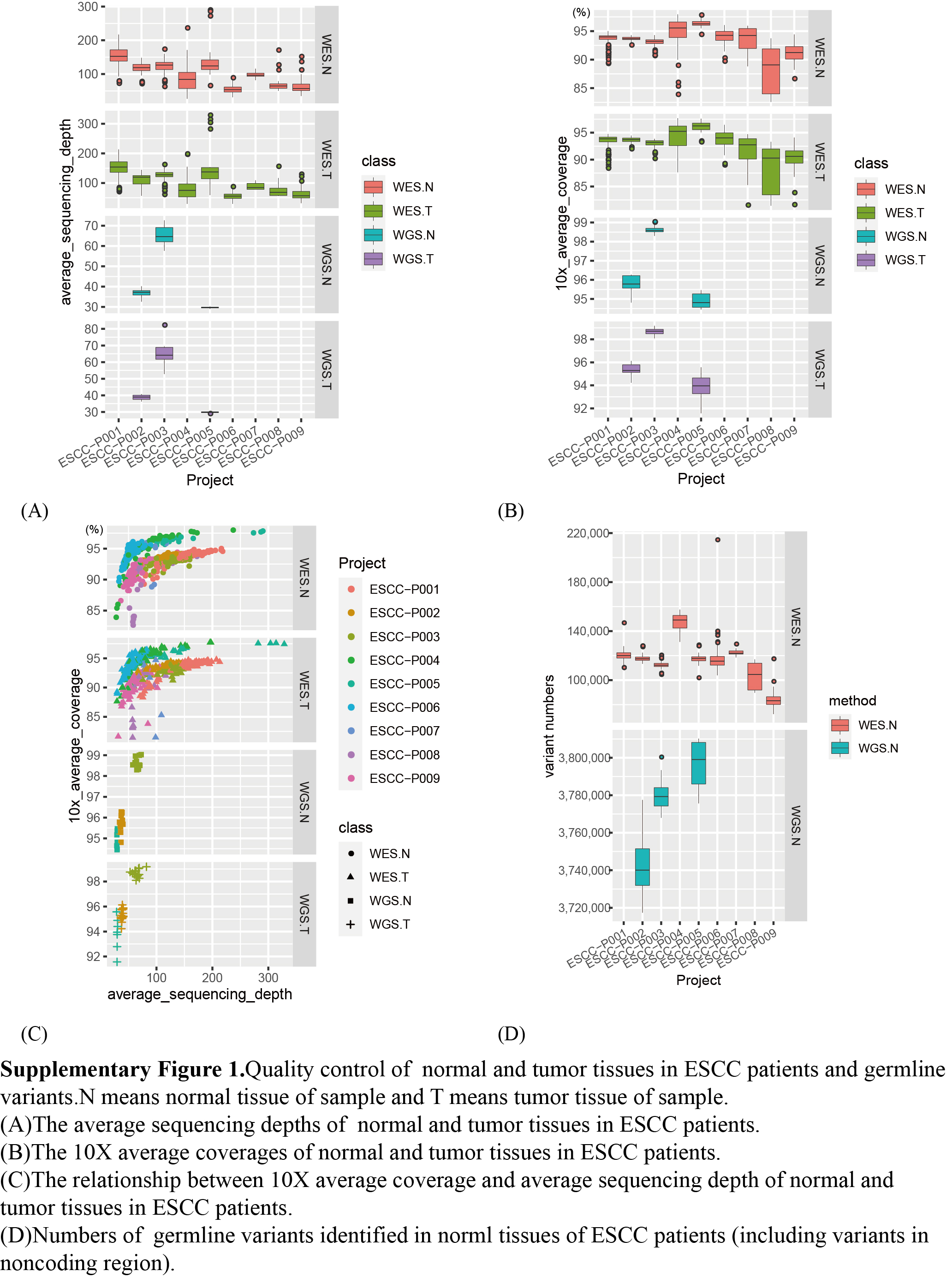

### The expression percentiles of TP53 and PTEN in ESCC-P006 cancer cohort

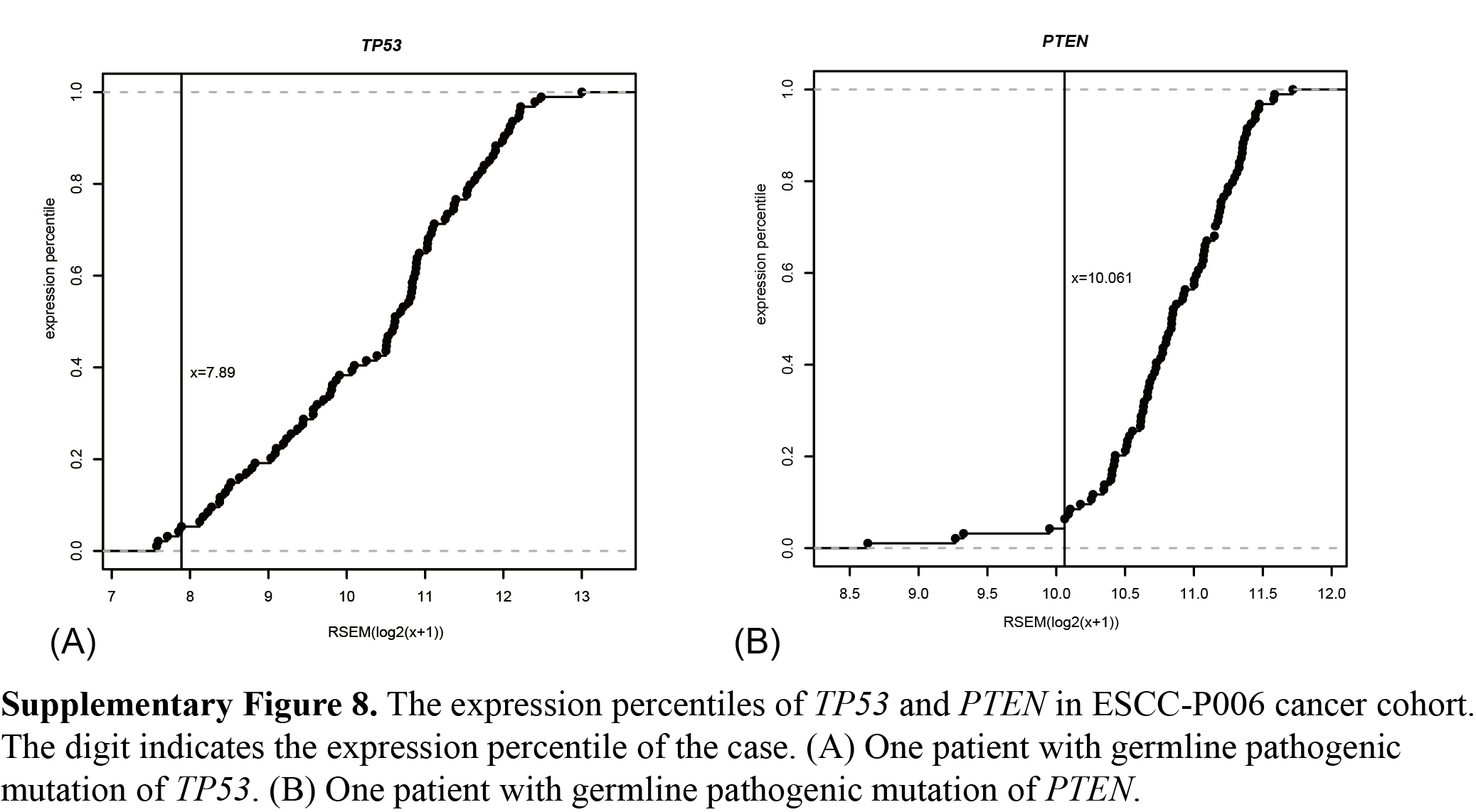

### The IGV screens showing germline mutations coupled with somatic mutations in ESCC samples

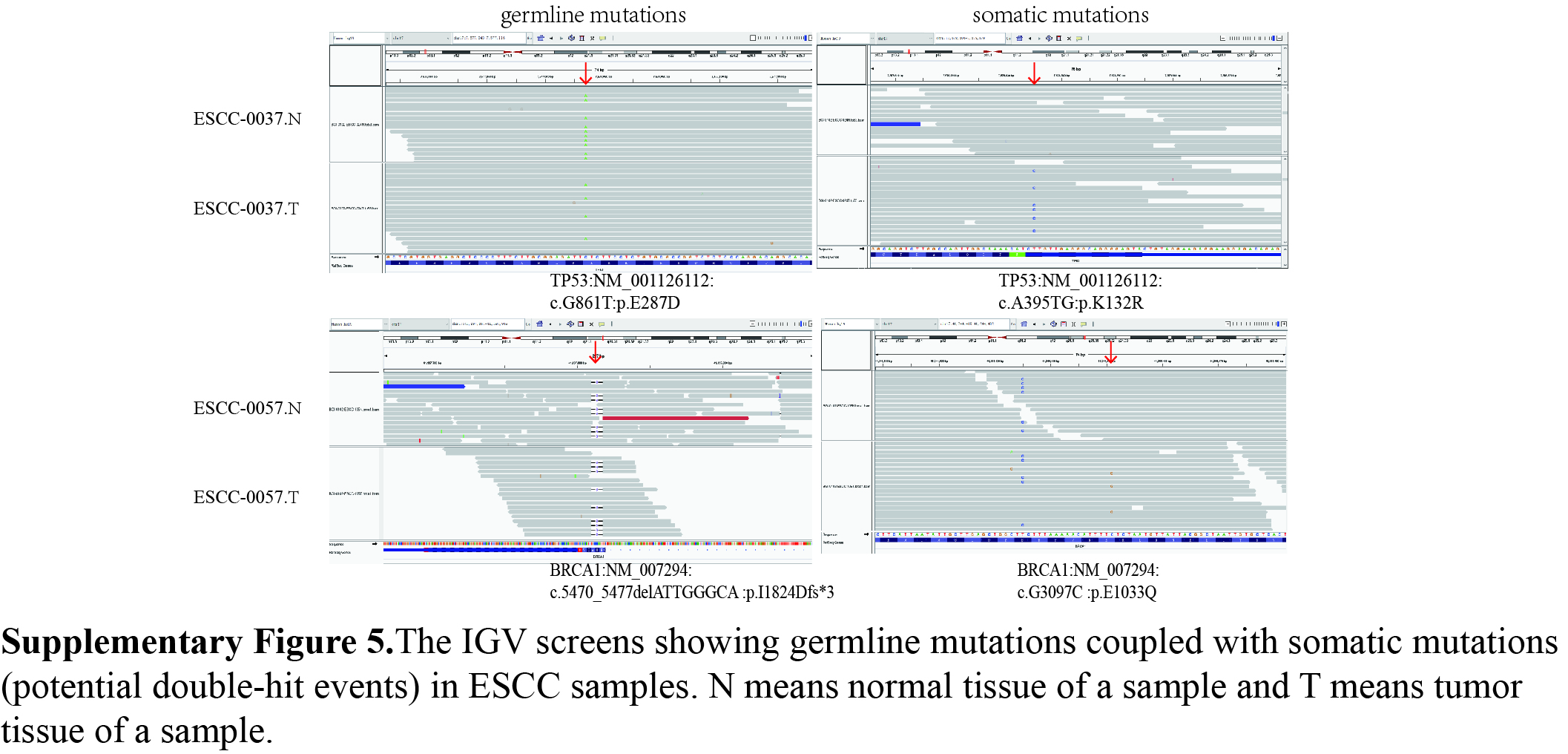

### The lollipop plot of pathogenic or likely pathogenic germline mutatinos in TP53 from 571 ESCC patients

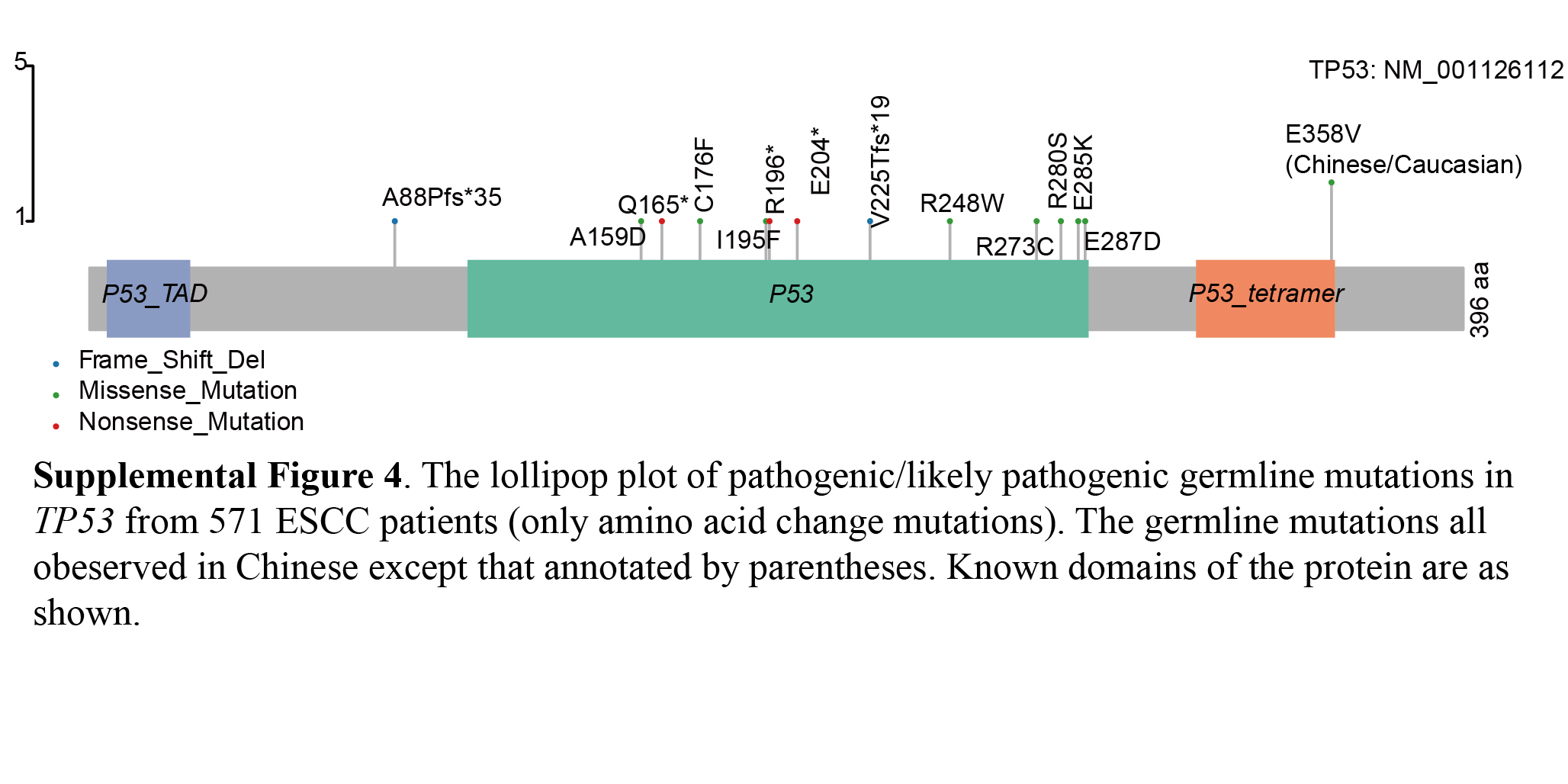

### The progress of how to select germline pathogenic variants

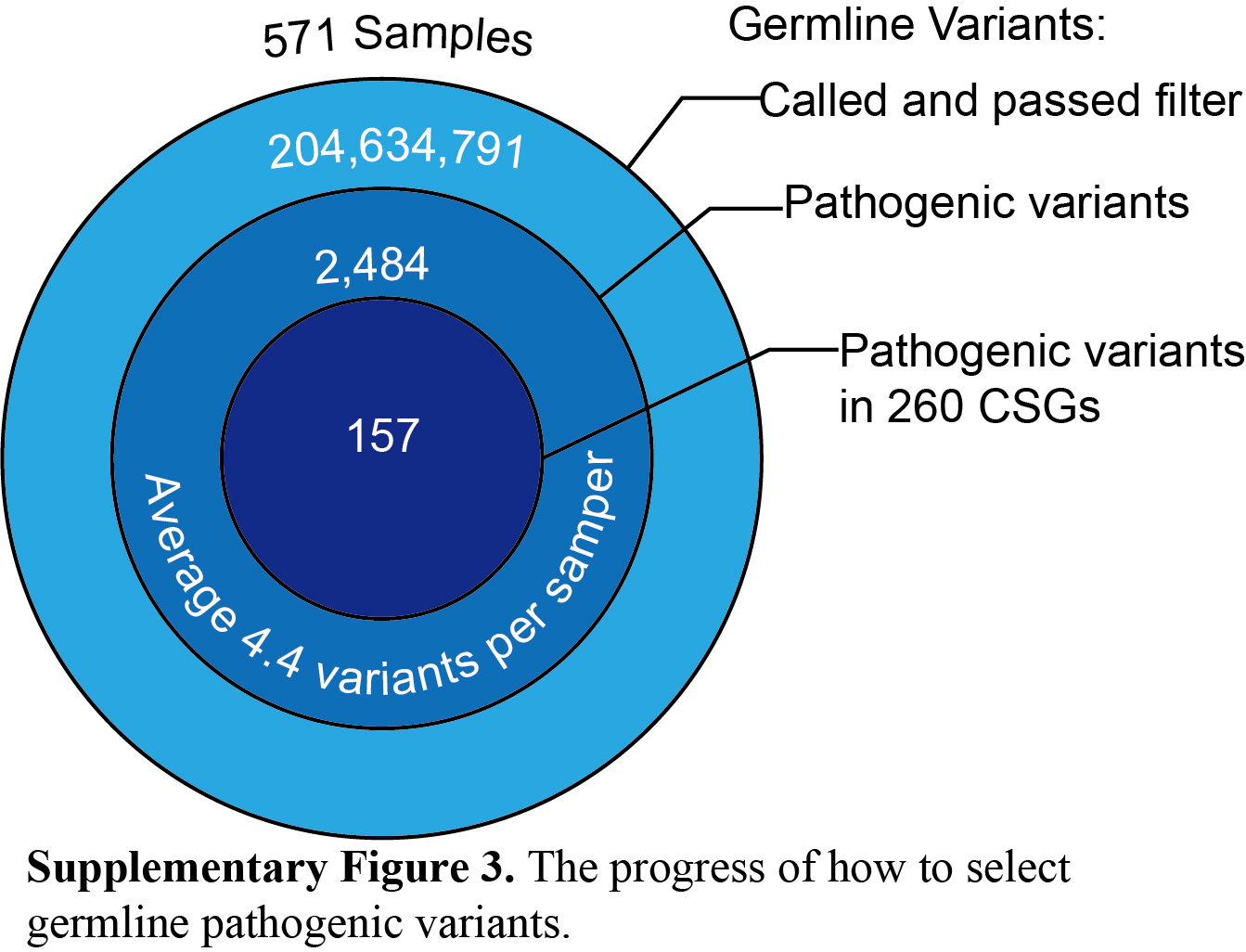

### The scatter plots showing germline mutations accompanied with allele loss SCNVs in normal tissues and tumor tissues of ESCC samples

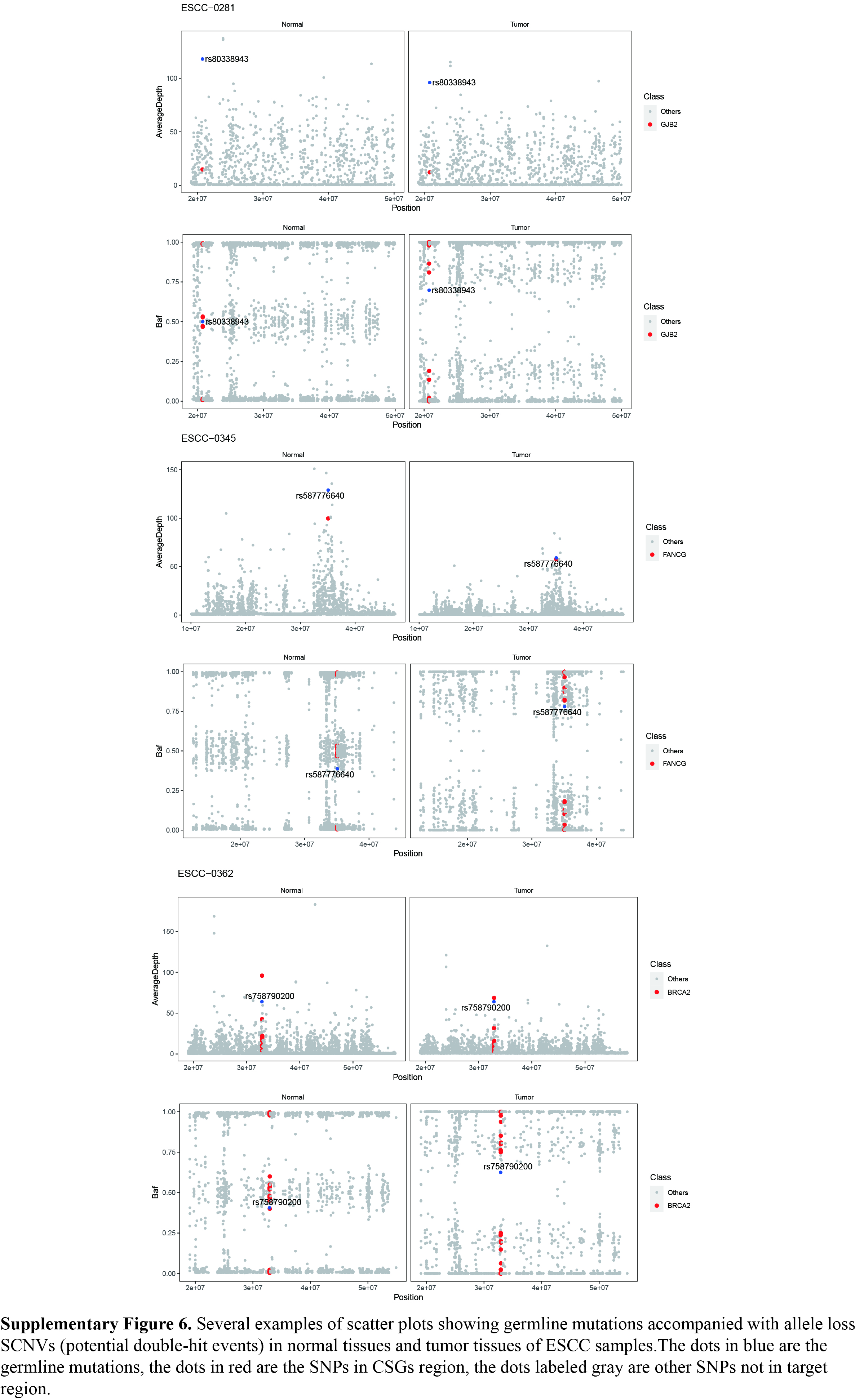

### The top 20 pathways of enrichment of four gene lists

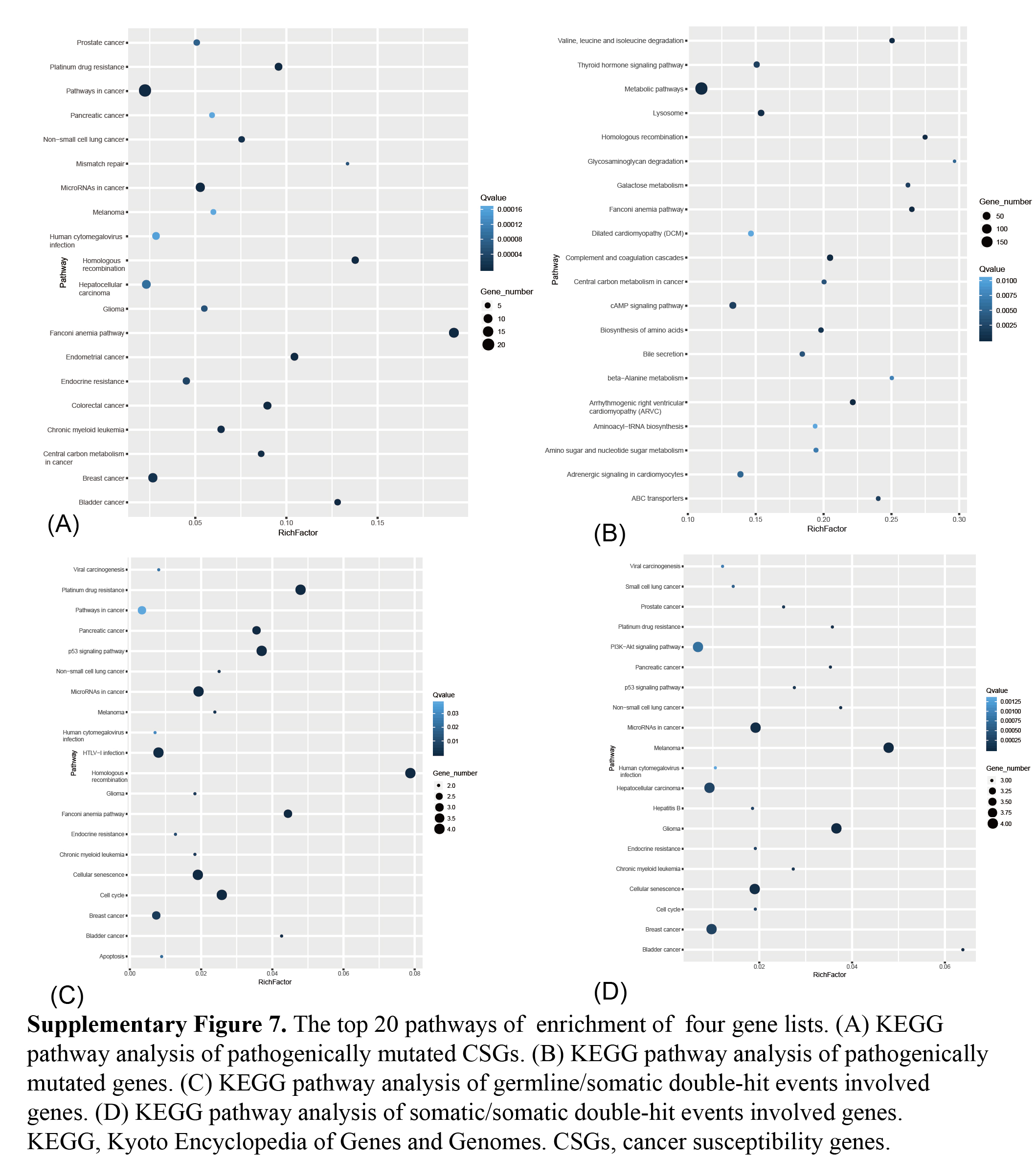
